# A RANDOMIZED CLINICAL TRIAL OF 2-WEEK METHOTREXATE DISCONTINUATION IN RHEUMATOID ARTHRITIS PATIENTS VACCINATED WITH INACTIVATED SARS-COV-2 VACCINE

**DOI:** 10.1101/2021.11.23.21266785

**Authors:** Carlo S R Araujo, Ana C Medeiros-Ribeiro, Carla G S Saad, Karina R Bonfiglioli, Diogo S Domiciano, Andrea Y Shimabuco, Matheus R Silva, Emily F N Yuki, Sandra G Pasoto, Tatiana N Pedrosa, Leonard Kupa, Gioanna Zou, Rosa M R Pereira, Clovis A Silva, Nadia E Aikawa, Eloisa Bonfa

## Abstract

**Objectives:** To evaluate the effect on immunogenicity and safety of 2-week methotrexate(MTX) discontinuation after each dose of the Sinovac-CoronaVac vaccine versus MTX maintenance in rheumatoid arthritis(RA) patients.

**Methods:** This was a single-center, prospective, randomized, investigator-blinded, intervention study (#NCT04754698, CoronavRheum), including adult RA patients(stable CDAI<10, prednisone<7.5mg/day), randomized(1:1) to withdraw MTX(MTX-hold) for 2 weeks after each vaccine dose or maintain MTX(MTX-maintain), evaluated at D0, D28 and D69. Co-primary outcomes were anti-SARS-CoV-2 S1/S2 IgG seroconversion(SC) and neutralizing antibody(NAb) positivity at D69. Secondary outcomes were GMT and changes in disease activity scores. For immunogenicity analyses, we excluded patients with baseline positive IgG/NAb, and, for safety reasons, those unable to hold MTX twice(CDAI>10 at D28).

**Results:** Randomization included 138 patients with 9 exclusions(5 COVID-19, 4 protocol violations). Safety evaluation included 60(MTX-hold) and 69(MTX-maintain) patients. Further exclusions consisted of 27 patients[13(21.7%) vs. 14(20.3%),p=0.848] with positive baseline IgG/NAb and 10 patients(21.3%) in MTX-hold with CDAI>10 at D28. At D69, a higher increase in SC[29(78.4%) vs 30(54.5%),p=0.019] was observed in MTX-hold(n=37) in comparison to MTX-maintain(n=55), with parallel augmentation in GMT[34.2(25.2-46.4) *vs* 16.8(11.9-23.6),p=0.006]. No differences were observed for NAb positivity[23(62.2%) *vs* 27(49.1%),p=0.217]. Longitudinal variations in disease activity scores were alike in both groups(CDAI,p=0.144; DAS28-CRP,p=0.718).

**Conclusion:** We provided novel data that 2-week MTX withdrawal after each vaccine dose improves anti-SARS-CoV-2 immunogenicity. The comparable longitudinal variations of disease activity in both groups suggest that discontinuation is a feasible and efficient strategy in well-controlled RA patients, and may be even safer for vaccines with longer interval between doses or single dose schedules.

**Funding:** FAPESP/CNPq/B3-Bolsa de Valores-Brasil.

## INTRODUCTION

The SARS-CoV-2 virus has caused a worldwide health, social and economic crisis with a death toll reaching millions,[1]. Brazil has been one of the most impacted countries, with mortality surpassing 600.000 subjects in October 2021,[2]. The World Health Organization (WHO) has recommended the emergency use of CoronaVac vaccine (Sinovac Life Sciences, Beijing,China),[3], an inactivated vaccine against SARS-CoV-2 CN02 strain, also one of the first approved vaccines in Brazil, accounting for over 70 million doses as of October 2021,[4]. The effectiveness of this vaccine was demonstrated in a large study with 10.2 million people in whom the protective effect for hospitalization, UCI admission and COVID-related death was over 85%,[5].

Rheumatoid arthritis (RA) patients are at higher risk of hospitalization and death by COVID-19 due to RA itself,[6], comorbidities,[7-8], or immunosuppressive treatments,[6-9]. Moreover, patients with RA have reduced immunogenicity to COVID-19 vaccines,[10-21], particularly those on glucocorticoids (GC),[10-13], methotrexate (MTX),[11-17], rituximab (RTX),[11-12, 17-21] and abatacept (ABA),[11-12, 21] therapies.

Temporary immunosuppressant withdrawal is suggested as a possible strategy to enhance vaccine immunogenicity in autoimmune rheumatic diseases (ARD) patients,[14-15, 22]. In this context, Park et al. demonstrated that the discontinuation of MTX improved immunogenicity of the annual influenza vaccines in RA patients,[14-15], concluding that the interruption of two MTX doses after vaccination was safe and equally effective as holding four MTX doses,[23-24]. Due to these results, recommendations have emerged favoring the withdrawal of MTX for 1-2 weeks after COVID vaccines,[25-26]. However, to this date, no comparative study has assessed the impact of this intervention on immunogenicity and disease activity after any SARS-CoV-2 vaccination schedule.

Therefore, this trial aimed to evaluate the immunogenicity and safety of 2-week MTX discontinuation after each dose of the Sinovac-CoronaVac vaccine in remission/low disease activity RA patients compared to age and sex balanced RA group who maintained the drug.

## METHODS

### Study design

This was a single-center, randomized, investigator-blind, intervention study performed at the rheumatology outpatient clinic of a tertiary center. The study was conducted in accordance with the Declaration of Helsinki and local regulations, and was approved by the institutional and national ethics committee (CAAE: 42566621.0.0000.0068). All RA patients fulfilled the American College of Rheumatology/European League Against Rheumatism criteria for the classification of RA,[27], and agreed to participate in the study and signed informed consents. The protocol is part of a larger study of immunosuppressed patients with ARD (Clinicaltrials.gov#NCT04754698),[12].

The co-primary outcomes were seroconversion (SC) rates for anti-SARS-CoV-2 S1/S2 IgG and neutralizing antibodies (NAb) positivity at D69. Secondary immunogenicity outcomes were: seroconversion rates for anti-S1/S2 IgG and NAb positivity at D28; geometric mean titer (GMT) and factor increase of GMT (FI-GMT) for anti-SARS-CoV-2 S1/S2 IgG and NAb activity at D28 and D69.

Secondary safety outcomes were longitudinal variations in disease activity scores: Clinical Disease Activity Index (CDAI),[28], Simplified Disease Activity Index (SDAI),[28], Disease Activity Score in 28 joints with C-reactive protein (DAS28-CRP),[29], and frequency of adverse events related to vaccine. Exploratory outcomes were the frequency of patients with flare at D28 and D69 defined by CDAI>10,[30, 31] or by an increase in DAS28-CRP >1.2 (or >0.6 if the baseline DAS28 was >3.2),[32-33]. Moreover, patient perception of disease activity worsening was also evaluated.

### Participants

We recruited adult (≥ 18 years old) patients with RA diagnosis [27] with low disease activity or remission (CDAI ≤10) [28] at first vaccination day and stable MTX dose for at least 4 weeks in monotherapy or in association with synthetic or biologic disease-modifying antirheumatic drugs (DMARD). Maximum allowed prevaccination oral prednisone dose was 7.5 mg/day. Patients were invited to participate after the review of their electronic records in the last 3 months (recruitment up to 3 weeks before enrollment). Exclusion criteria were acute febrile illness/symptoms of COVID-19 at vaccination, history of anaphylaxis to vaccine components, demyelinating disease, decompensated heart failure (class III/IV), blood transfusion ≤6 months, inactivated virus vaccine ≤14 days, live virus vaccine <4 weeks, denial to participate, hospitalization, previous vaccination with any SARS-CoV-2 vaccine, reverse-transcriptase-polymerase-chain-reaction (RT-PCR) confirmed COVID-19 during the study, and rituximab use in the previous 12 months. Patients with prevaccination positive COVID-19 serology (anti-S1/S2 IgG and/or NAb) were excluded from immunogenicity analysis but kept for safety evaluation.

### Visit schedule

Patients were evaluated in three visits: D0 (first dose of the vaccine), D28 (second dose) and D69 (6 weeks after the 2^nd^ dose). First dose was given on February, 9^th^-10^th^, 2021 (D0), while the second dose was on March 9-10^th^, 2021 (D28).

The vaccination protocol included two doses of ready-to-use syringes with Sinovac-CoronaVac vaccine (Sinovac Life Sciences, Beijing, China, batch #20200412), consisting of 3 µg in 0.5 mL of β-propiolactone inactivated SARS-CoV-2 with aluminum hydroxide as an adjuvant. Vaccine was administered intramuscularly in the deltoid area.

### Randomization and masking

Investigators responsible for disease activity measures, statisticians and laboratory personnel were blinded to the allocation groups. Only 2 researchers (C.S.R.A. and M.R.S.) were not blinded and were responsible for safety surveillance and patient follow-up by telephone for adherence purposes. These 2 investigators were not involved in disease activity measures, laboratory analysis or patient vaccination.

At D0, before the first vaccine dose, patients were evaluated by blinded experienced rheumatologists who assessed disease activity by CDAI and rechecked inclusion and exclusion criteria. Patients with CDAI<10 proceeded to the enrollment station, where the unblinded researchers revised the protocol, explained the procedures, collected the informed consent and conducted the randomization, which was performed on the web-based software “The REDcap Project version 10.5.0”. Allocation was generated instantaneously in a 1:1 ratio to one of the following groups: withdraw MTX for 2 weeks after each dose of the CoronaVac (MTX-hold group) or to maintain MTX continuously (MTX-maintain group).

At D28 and D69, patients were initially assessed by the unblinded researchers, checked for protocol violation, and instructed not to inform their allocation groups to anyone else. Then, they proceeded for the blinded disease activity evaluation. Subsequently, they returned to the unblinded researchers and were instructed accordingly.

### Intervention

At D0, the two unblinded researchers instructed patients in the MTX-hold group not to take 2 doses of MTX after vaccination, according to the last MTX dose. They provided a date diagram (Supplementary figure 1) informing the dates in which they would skip MTX, and the date to resume its usage. At D28, patients in the MTX-hold group with CDAI<10 were instructed to withdraw MTX again, and a new date diagram was produced. Patients with CDAI>10 in the MTX-hold group were instructed not to withdraw MTX again after vaccine 2^nd^ dose. Patients in the MTX-maintain group were instructed to continue MTX on the same day and dose throughout the study. The 2 unblinded researchers checked adherence to protocol by telephone contact with all patients in the weeks following both vaccine doses.

Adding or changing DMARD therapy was not allowed until D69, though patients were permitted to use analgesics, non-steroidal anti-inflammatory drugs or prednisone up to 10 mg/day in case of disease activity worsening.

### Laboratory analyses

Blood samples (30 mL) were collected immediately before each vaccine dose (D0 and D28) and 6 weeks after the 2^nd^ dose (D69). Serum samples were stored at -70 °C. IgG antibodies against the SARS-CoV-2 S1/S2 proteins were measured using a chemiluminescent immunoassay (Indirect ELISA, LIAISON®, DiaSorin, Italy). SC was defined as positive serology (≥ 15.0 UA/mL). GMT and 95% confidence intervals were calculated attributing the value of 1.9 UA/mL to undetectable levels (<3.8 UA/mL). FI-GMT was calculated as the ratio of the IgG titer after vaccination to the IgG titer before vaccination. Detection of NAb was performed using the SARS-CoV-2 sVNT Kit (GenScript, Piscataway, NJ, USA). Positivity was defined as ≥30% inhibition of this linkage,[34]. Medians (interquartile range) of the percentage of neutralizing activity were only calculated for positive samples. CRP (by the nephelometric method), was also measured.

### Safety outcomes

Disease activity was checked by experienced rheumatologists, blinded to allocation groups, who assessed the following parameters: number of tender joints, number of swollen joints (both in 28 joint count), patient global assessment of disease activity (PGA, by visual analog scale - VAS) and evaluator global assessment of disease activity (EGA - by VAS). With this data, CDAI was calculated. With CRP from sera collected on the same day, SDAI and DAS28-CRP were also calculated.

Patients were instructed to fill a structured diary of local (pain, erythema, swelling, bruise, pruritus, and induration at the vaccine site) and systemic symptoms (fever, malaise, somnolence, lack of appetite, nausea, vomit, diarrhea, abdominal pain, vertigo, tremor, headache, fatigue, myalgia, muscle weakness, arthralgia, back pain, cough, sneezing, coryza, stuffy nose, sore throat, shortness of breath, conjunctivitis, pruritus, and skin rash) after each vaccination to explore potential vaccine side effects. Adverse effect severity was classified according to WHO definition, [35]. Patients who had symptoms suggestive of COVID-19 infection had nasopharyngeal RT-PCR tests done.

### Statistical analysis

The sample size calculation was based on the 2009 non-adjuvanted influenza A/H1N1 primo vaccination in a large cohort of RA patients under MTX, which induced SC rate of 46%, [36]. Expecting an increment of 20% in the MTX-hold group, [14-15], that should achieve 66% SC rate, with a 5% alpha error and 80% power (1:1 ratio), the minimum sample would be of 96 patients per group.

Categorical variables were presented as number (percentage) and compared using chi-square or Fisher’s exact tests, as appropriated. Continuous general data were presented as medians (interquartile ranges) and compared using t-test or Mann-Whitney test, as appropriate. Data regarding IgG titers and disease activity scores at different time points were analyzed using generalized estimating equations (GEE) with normal marginal distribution and gamma distribution respectively and identity binding function assuming first order autoregressive correlation matrix between moments (D0, D28 and D69) in the comparison of the 2 groups (MTX-hold and MTX-maintain), followed by Bonferroni’s multiple comparisons. IgG titers were analyzed as neperian logarithm (ln)-transformed data. Multiple regression analyses were performed including SC or presence of NAb at D69 as the dependent variables and as independent variables those with p<0.2 in the univariate analyses. Statistical significance was defined as p<0.05. All statistical analyses were performed using Statistical Package for the Social Sciences, version 20.0 (IBM-SPSS for Windows. 20.0. Chicago, IL, USA).

## RESULTS

A total of 247 RA patients fulfilled the profile on electronic chart review and were pre-selected. After exclusion criteria, 138 patients remained and were randomized, with 67 in the MTX-hold and 71 in the MTX-maintain groups (Figure 1). During the study, there were 9 further exclusions: 5 RT-PCR-confirmed COVID-19 and 4 protocol violations. Therefore, final groups for all safety analysis and disease activity evaluation consisted of 60 patients in the MTX-hold and 69 in the MTX-maintain groups. For the immunogenicity analysis, patients with positive anti-SARS-CoV-2 serology/NAb [13(21.7%) *vs* 14(20.3%), respectively, p=0.848] at D0 were excluded and groups consisted of 47 (MTX-hold) *vs* 55 patients (MTX-maintain). Out of the 47 patients in the MTX-hold group, 10 (21.3%) had a flare on D28 and did not stop MTX the second time, thus 37 patients comprehended the MTX-hold group for immunogenicity and finished the complete MTX withdrawal protocol. MTX-hold and MTX-maintain groups had similar age and female sex frequencies (p>0.05). Other demographic characteristics, comorbidities, disease duration, baseline disease activity, rheumatoid factor and anti-CCP positivity, current therapy did not differ between the two groups (p>0.05), except for the association with prednisone which was more frequent in the MTX-hold group regarding safety analyses (p=0.021) (Table 1).

**Figure 1.**
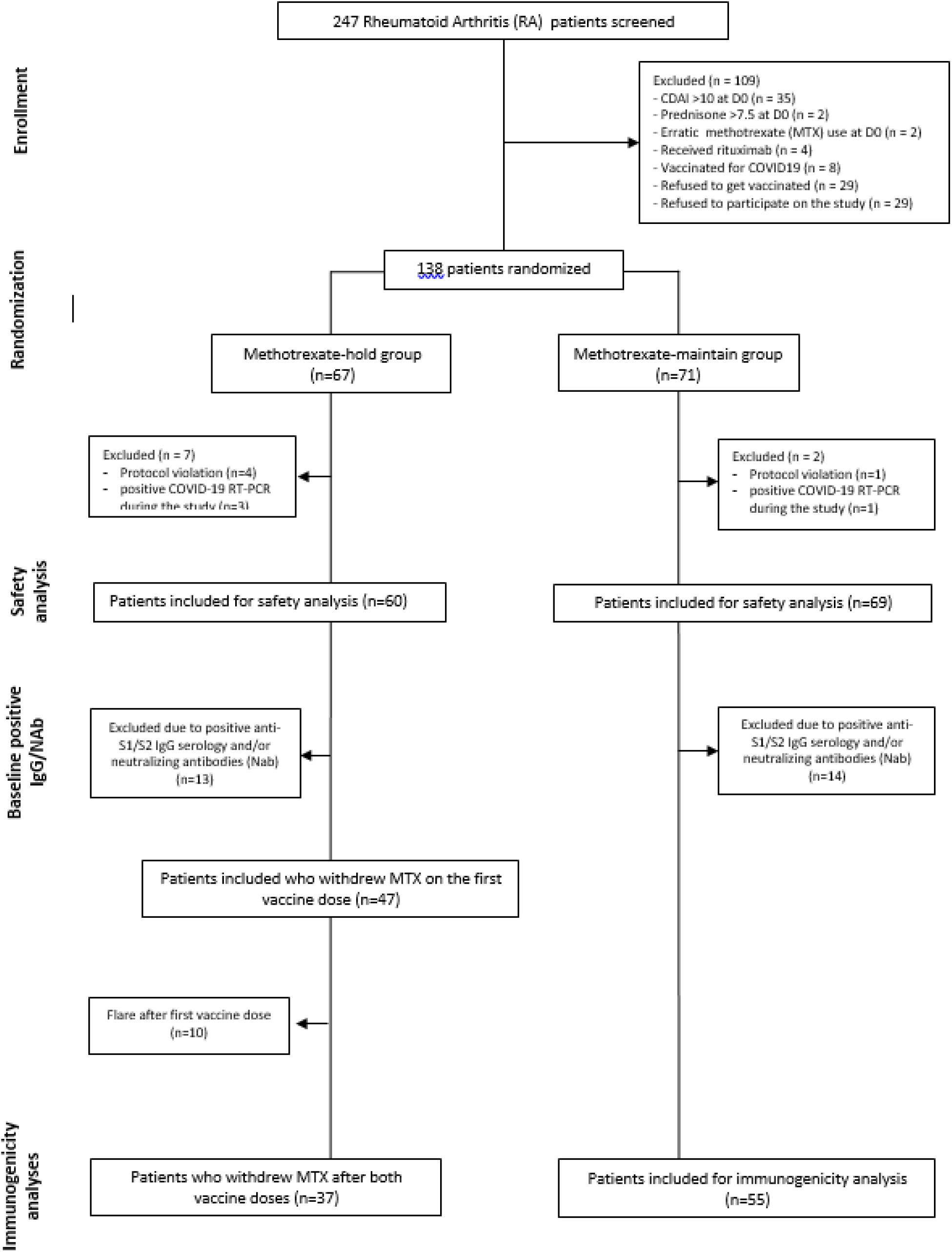
Modified CONSORT flow diagram.

**Table 1.** Baseline characteristics in rheumatoid arthritis (RA) patients according to methotrexate (MTX) interruption (MTX-hold) or maintenance (MTX-maintain)

|  | Patients for safety analyses |  |  | Patients for immunogenicity analyses |  |  |
| --- | --- | --- | --- | --- | --- | --- |
|  | MTX-hold<br>(n=60) | MTX-maintain<br>(n=69) | *p* | MTX-hold<br>(n=37) | MTX-maintain<br>(n=55) | *p* |
| Current age, years | 61 (52-70.5) | 58 (49-68) | 0.395 | 59 (45-68) | 59 (51.5-68) | 0.828 |
| Female sex | 51 (85.0) | 63 (91.3) | 0.265 | 34 (91.9) | 50 (90.9) | >0.999 |
| Caucasian race | 28 (46.7) | 29 (42) | 0.597 | 19 (51.4) | 25 (45.5) | 0.579 |
| Disease parameters |  |  |  |  |  |  |
| Disease duration | 17(10-30.5) | 17 (10-24) | 0.801 | 19 (10-28) | 17(10-25) | 0.616 |
| RF positivity | 48 (80) | 53/68*(77.9) | 0.776 | 31 (83.8) | 44(80) | 0.647 |
| Anti-CCP positivity | 27/37* (73) | 35/47* (74.5) | 0.877 | 17/22* (77.3) | 26/36* (72.2) | 0.670 |
| CRP, mg/dl | 4.4 (1.1-9.3) | 3.7 (1.6-7.0) | 0.655 | 5.6 (1.0-9.6) | 4.0 (2.5-7.5) | 0.847 |
| CDAI | 7.0 (4.0-9.0) | 6.0 (3.0-8.0) | 0.485 | 6.0 (2.0-8.0) | 6.0 (3.0-8.0) | 0.548 |
| SDAI | 7.1 (4.1-10.1) | 6.3 (3.9-9.3) | 0.380 | 6.8 (3.1-9.4) | 6 (3.8-8.3) | 0.618 |
| DAS28-CRP | 2.72 (2.10-3.15) | 2.33 (1.96-2.82) | 0.178 | 2.43 (1.80-3.15) | 2.34 (2.02-2.87) | 0.927 |
| CDAI≤2.8 | 11 (18.3) | 10 (14.5) | 0.556 | 10 (27) | 8 (14.5) | 0.139 |
| SDAI≤3.3 | 12 (20) | 14 (20.3) | 0.967 | 12 (32.4) | 11 (20) | 0.177 |
| DAS28-CRP≤2.6 | 30 (50) | 46 (66.6) | 0.055 | 21 (56.8) | 36 (65.5) | 0.400 |
| Boolean criteria | 9 (15) | 14 (20.3) | 0.434 | 9 (24.3) | 12 (21.8) | 0.779 |
| Current therapy |  |  |  |  |  |  |
| Prednisone | 33 (55) | 24 (34.8) | 0.021 | 19 (51.4) | 18 (32.9) | 0.074 |
| Prednisone dose, mg/dl | 5 (4.38-5) | 5 (5-5) | 0.495 | 5 (2.5-5) | 5 (5-5) | 0.336 |
| MTX dose | 20 (17.5-25) | 20 (15-25) | 0.397 | 20 (15-25) | 20 (15-25) | 0.314 |
| MTX 10-15 mg/week | 16 (26.7) | 23 (33.3) | 0.411 | 11 (29.7) | 20 (36.4) | 0.509 |
| MTX 17.5-25 mg/week | 44 (73.3) | 46 (66.7) |  | 26 (70.3) | 35 (63.6) |  |
| MTX monotherapy | 13 (21.7) | 18 (26.1) | 0.558 | 9 (24.3) | 16 (29.1) | 0.614 |
| Leflunomide | 8 (13.3) | 17 (24.6) | 0.105 | 6 (16.2) | 13 (23.6) | 0.389 |
| Sulfasalazine | 1 (1.7) | 0 | 0.465 | 1 (2.7) | 0 | 0.402 |
| Hydroxychloroquine | 7 (11.7) | 16 (23.2) | 0.088 | 4 (10.8) | 13 (23.6) | 0.172 |
| Tofacitinib | 2 (3.3) | 3 (4.3) | >0.999 | 1 (2.7) | 3 (5.5) | 0.646 |
| TNF inhibitor | 14 (23.3) | 11 (15.9) | 0.290 | 10 (27) | 8 (14.5) | 0.139 |
| Abatacept | 7 (11.7) | 9 (13) | 0.813 | 6 (16.2) | 7 (12.7) | 0.638 |
| Tocilizumab | 4 (6.7) | 3 (4.3) | 0.704 | 1 (2.7) | 1 (1.8) | >0.999 |

Results are expressed in median (interquartile ranges) and n (%). Continuous data are compared using the Mann–Whitney U-test, and categorical variables with the chi-square or Fisher’s exact test, as appropriate, as two-sided analyses. For safety analyses, all patients who adhered to protocol were included. For immunogenicity analyses, patients with baseline positive S1/S2 IgG were excluded from both groups, and patients with CDAI>10 at D28 that withdrew MTX only once were excluded only from the MTX-hold group; \* - indicates that percentages calculated according to available data. MTX- methotrexate; RF - rheumatoid factor; anti-CCP - anti-cyclic citrullinated peptides; CDAI - clinical disease activity index; SDAI - simplified disease activity index; DAS28-CRP - disease activity score with 28 joints and C-reactive protein; TNF - tumor necrosis factor.

### Immunogenicity outcomes

Baseline anti-SARS-CoV-2 S1-S2 IgG GMT [2.1(1.9-2.3) vs 2.0(1.9-2.1), p>0.999] were similar among groups. At D69, patients who withdrew MTX after both vaccine shots (MTX-hold, n=37) had higher SC [29(78.4%) *vs*. 30(54.5%), p=0.019], with a parallel augmentation in GMT [34.2(25.2-46.4) *vs*. 16.8(11.9-23.6), p=0.006] and a higher FI-GMT [17.1(12.6-23.1) *vs*. 8.1(5.8-11.4), p=0.007] in comparison to MTX-maintain group (n=55) (Table 2 and Figure 2). For NAb positivity, the difference was not significant [23(62.2%) *vs*. 27(49.1%), p=0.217], as also occurred for NAb activity [53%(42-68.8) vs. 51.7%(37.8-62.2), p=0.335] (Table 3).

**Table 2.** Data regarding anti-S1/S2 IgG [seroconversion (SC) rates, anti-SARS-CoV-2 S1/S2 IgG titers and Factor Increase (FI) in titers], after the first and second doses of Sinovac-CoronaVac vaccine in rheumatoid arthritis (RA) patients according to methotrexate (MTX) interruption (MTX-hold) or maintenance (MTX-maintain)

|  | After 1<sup>st</sup><br>dose (D28) | After 2<sup>nd</sup><br>dose (D69) | Baseline<br>(D0) | After 1<sup>st</sup><br>dose (D28) | After 2<sup>nd</sup><br>dose (D69) | From D0 to<br>D28 | From D0 to<br>D69 |
| --- | --- | --- | --- | --- | --- | --- | --- |
|  | SC | SC | GMT | GMT | GMT | FI-GMT | FI-GMT |
| MTX-hold<br>(n=37) | 10<br>(27.0) | 29<br>(78.4) | 2.1<br>(1.9-2.3) | 6.8*<br>(4.8-9.6) | 34.2*<br>(25.2-46.4) | 3.4<br>(2.4-4.7) | 17.1<br>(12.6-23.1) |
| MTX-maintain<br>(n=55) | 2<br>(3.6) | 30<br>(54.5) | 2.0<br>(1.9-2.1) | 2.8<br>(2.3-3.5) | 16.8*<br>(11.9-23.6) | 1.4<br>(1.1-1.7) | 8.1<br>(5.8-11.4) |
| p | 0.003 | 0.019 | >0.999 | 0.002 | 0.006 | <0.001 | 0.007 |

**Table 3.** Neutralizing antibodies (NAb) and neutralizing activity, after the first and second doses of Sinovac-CoronaVac vaccine in rheumatoid arthritis (RA) patients according to methotrexate (MTX) interruption (MTX-hold) or maintenance (MTX-maintain)

|  | After vaccine 1<sup>st</sup> dose |  | After vaccine 2<sup>nd</sup> dose |  |
| --- | --- | --- | --- | --- |
|  | % patients with positive NAb | Median neutralizing activity in NAb positive patients (%) | % patients with positive NAb | Median neutralizing activity in NAb positive patients (%) |
| MTX-hold (n=37) | 8<br>(21.6) | 41<br>(34.8-49.7) | 23<br>(62.2) | 53<br>(42-68.8) |
| MTX-maintain (n=55) | 4<br>(7.3) | 57.7<br>(51.8-65.9) | 27<br>(49.1) | 51.7<br>(37.8-62.2) |
| P | 0.057 | 0.345 | 0.217 | 0.335 |

MTX-hold - baseline seronegative patients randomized to interrupt MTX after the 1<sup>st</sup> and 2<sup>nd</sup> doses, excluding those who had CDAI>10 at D28 and withdrew MTX only once. MTX-maintain - baseline seronegative patients randomized to maintain methotrexate throughout the study and who adhered to the protocol. Frequencies of subjects with positive NAb are expressed as number (%) and were compared using a chi-square or Fisher's exact test, as appropriate. Positivity for NAb was defined as neutralizing activity ≥ 30% (cPass sVNT Kit). Percentages of neutralizing activity among subjects with positive NAb are expressed as median (interquartile range). Data were compared using a two-sided Mann-Whitney U-test for comparison between MTX-hold and MTX-maintain groups at prespecified time points (D28 and D69).

**Figure 2.**
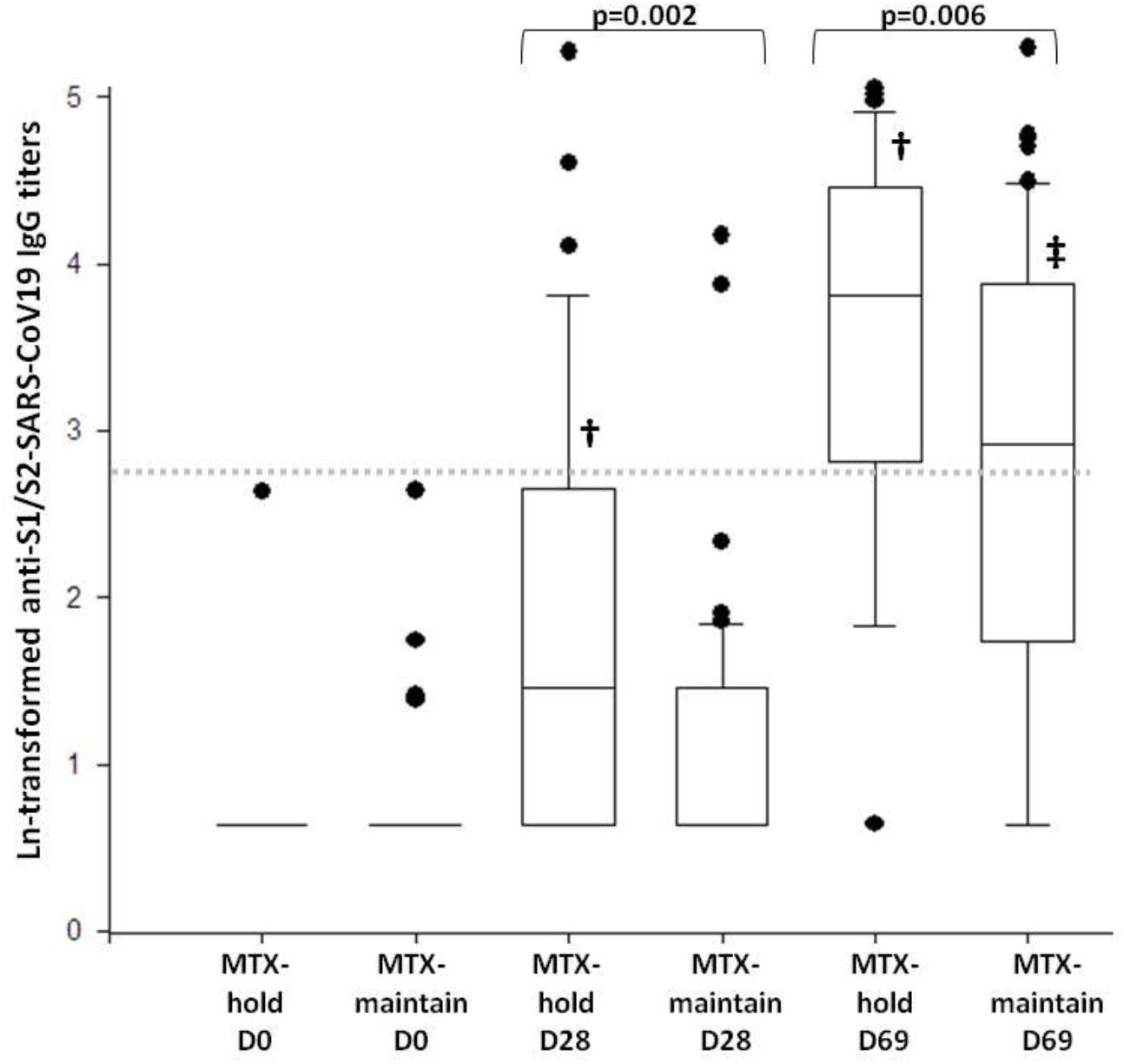
Box plots of anti-S1/S2 IgG titers at baseline and after the first and second doses of Sinovac-CoronaVac vaccine in rheumatoid arthritis (RA) patients according to methotrexate (MTX) interruption (MTX-hold) or maintenance (MTX-maintain) MTX-hold - baseline seronegative patients randomized to interrupt MTX after the 1^st^ and 2^nd^ doses, excluding those who had CDAI>10 at D28 and withdrew MTX only once. MTX-maintain - baseline seronegative patients randomized to maintain methotrexate throughout the study and who adhered to the protocol. Analyses were performed with neperian logarithm (ln)-transformed data using generalized estimating equations (GEE) with normal marginal distribution and gamma distribution respectively, and identity binding function assuming first order autoregressive correlation matrix between moments (D0, D28 and D69) in the comparison of the 2 groups (MTX-hold and MTX-maintain), followed by Bonferroni’s multiple comparisons. The mean behavior of the ln-transformed IgG titers was different in MTX-hold and MTX-maintain (p interaction=0.001). Groups were comparable at baseline (p>0.999), but MTX-hold group had higher mean titers at D28 (p=0.002) and at D69 (p=0.006). Mean titers increased at each time point for MTX-hold group (†p<0.001 from D0 to D28 and from D28 to D69). For MTX-maintain group, the titers did not increase from D0 to D28 (p=0.423) but increased from D28 to D69 (‡p<0.001). All analyses were two-sided. Dotted line denotes the cut-off level for positivity (ln 15 AU ml–1 = 2.71 by Indirect ELISA, LIAISON SARS-CoV-2 S1/S2 IgG).

In a further analysis combining both groups, the comparison of patients who had SC and those who did not seroconvert showed that older age, age > 60 years and combination with LEF were negatively associated with SC, while MTX withdrawal twice was positively related to it. For NAb, only older age and age > 60 years were negatively associated with absence of NAb (Supplemental Table 1). In multivariate analyses, older age [OR 0.71 (0.56-0.89) for each 5-year interval, p=0.003] and age > 60 years [OR 0.16 (0.05-0.50), p=0.001] persisted negatively associated with SC, while MTX withdrawal twice [OR 4.6 (1.43-15.04), p=0.010] was positively associated with SC.

### Disease activity

For disease activity evaluation, the groups consisted of 60 (MTX-hold) and 69 (MTX-maintain) patients, including those with positive baseline IgG/NAb. Longitudinally, CDAI (p=0.144), SDAI (p=0.117), DAS28-CRP (p=0.718), and CRP (p=0.410) had the same behavior in MTX-hold and MTX-maintain groups, with worsening at D28, but not from D28 to D69 (Figure 3).

**Figure 3.**
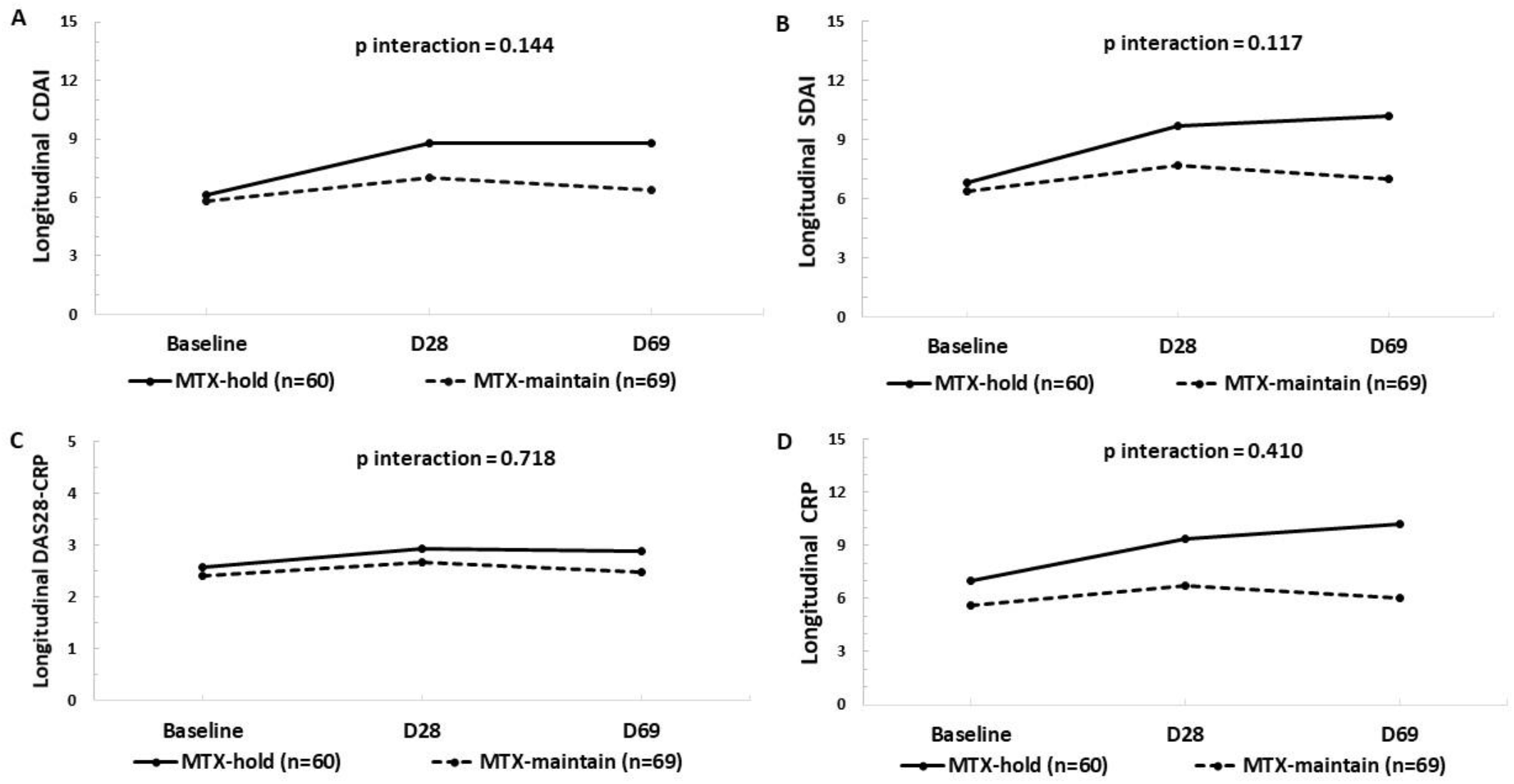
Analyses of continuous disease activity parameters at baseline, after the first and second doses of Sinovac-CoronaVac vaccine in rheumatoid arthritis (RA) patients according to methotrexate (MTX) interruption (MTX-hold) or maintenance (MTX-maintain) MTX-hold - baseline seronegative patients randomized to interrupt MTX after the 1^st^ and 2^nd^ doses, excluding those who had CDAI>10 at D28 and withdrew MTX only once. MTX-maintain - baseline seronegative patients randomized to maintain MTX throughout the study and who adhered to the protocol. Data regarding disease activity parameters are shown as means and were analyzed using generalized estimating equations (GEE) with normal marginal distribution and gamma distribution respectively, and identity binding function assuming first order autoregressive correlation matrix between moments (D0, D28 and D69) in the comparison of the 2 groups (MTX-maintain and MTX-hold), followed by Bonferroni’s multiple comparisons. CDAI - clinical disease activity index; SDAI - simplified disease activity index; DAS28-CRP - disease activity score with 28 joints and C-reactive protein; CRP - C-reactive protein. The mean behavior of CDAI (Figure 3A), SDAI (Figure 3B), DAS28-CRP (Figure 3C), and CRP (Figure 3D) was similar in MTX-hold and MTX-maintain throughout the study (p=0.144, p=0.117, p=0.718, and p=0.410, respectively), increasing after the first dose (p<0.001, p<0.001, p<0.001, and p=0.021, respectively) and remaining stable after the second dose (p>0.999, p>0.999, p=0.602, and p>0.999, respectively).

At D28, no differences appeared regardless of the flare definition (p>0.05) (Table 4). At D69, both groups had similar rates of flares based on DAS28 variations [12(20%) *vs*. 8(11.6%), p=0.188] (Table 4). However, MTX-hold group had a greater number of patients who lost their initial condition based on CDAI [19(31.7%) *vs*. 9(13%), p=0.011] and patients reported a higher perception of disease worsening [8(13.3%) *vs*. 2(2.3%), p=0.044] (Table 4). The magnitude of variation of CDAI among patients who flared was similar between groups [9(4-13.5) *vs*. 7(4.8-10.8), p=0.456].

**Table 4.**
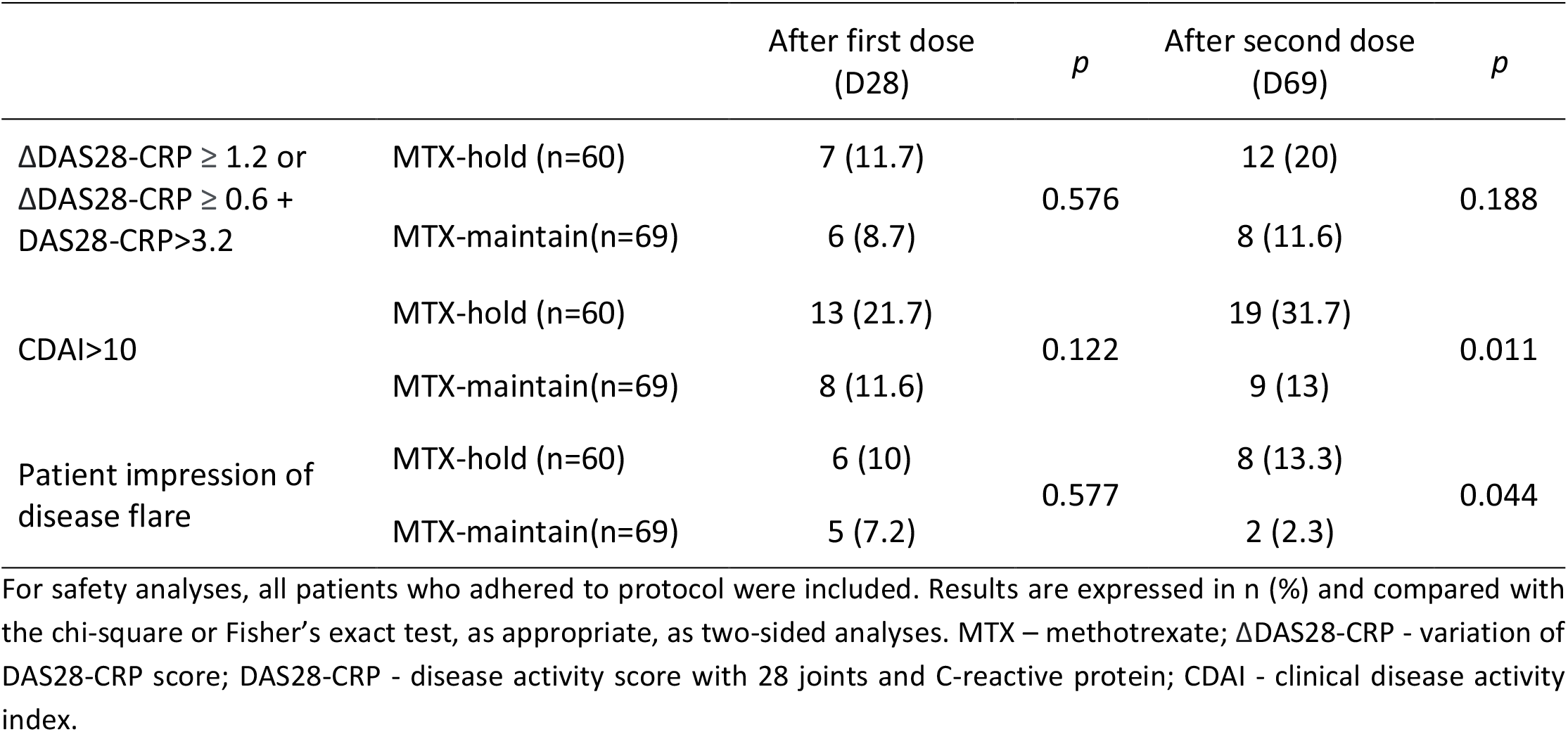
Disease activity analyses after the first and second doses of Sinovac-CoronaVac vaccine in rheumatoid arthritis (RA) patients.

|  |  | After first dose<br>(D28) | *p* | After second dose<br>(D69) | *p* |
| --- | --- | --- | --- | --- | --- |
| ΔDAS28-CRP $\geq 1.2$ or<br>ΔDAS28-CRP $\geq 0.6$ +<br>DAS28-CRP $> 3.2$ | MTX-hold (n=60) | 7 (11.7) | 0.576 | 12 (20) | 0.188 |
|  | MTX-maintain(n=69) | 6 (8.7) |  | 8 (11.6) |  |
| CDAI $> 10$ | MTX-hold (n=60) | 13 (21.7) | 0.122 | 19 (31.7) | 0.011 |
|  | MTX-maintain(n=69) | 8 (11.6) |  | 9 (13) |  |
| Patient impression of<br>disease flare | MTX-hold (n=60) | 6 (10) | 0.577 | 8 (13.3) | 0.044 |
|  | MTX-maintain(n=69) | 5 (7.2) |  | 2 (2.3) |  |

For safety analyses, all patients who adhered to protocol were included. Results are expressed in n (%) and compared with the chi-square or Fisher's exact test, as appropriate, as two-sided analyses. MTX – methotrexate; $\Delta$ DAS28-CRP - variation of DAS28-CRP score; DAS28-CRP - disease activity score with 28 joints and C-reactive protein; CDAI - clinical disease activity index.

### Vaccine side effects

Approximately half of the patients reported mild side effects, without differences between the groups. After the 2^nd^ vaccine dose, myalgia [10(16.7%) vs. 3(4.3%), p=0.037] and vertigo [7(11.7%) vs. 1(1.5%), p=0.024] were more frequent in the MTX-hold group (Supplemental Table 2).

## DISCUSSION

To the best of our knowledge, this is the first randomized study to compare the impact of MTX withdrawal in the immunogenicity and disease activity of any COVID-19 vaccine in RA patients. We demonstrated that temporary suspension is effective in increasing IgG SC and GMT levels. The observed comparable disease activity variation in MTX-hold and MTX-maintain groups suggests that this strategy is effective and safe to boost vaccine immunogenicity in well-controlled RA patients under MTX therapy.

The study has some strengths such as the inclusion of patients in remission/low disease activity and low prednisone doses, providing a safer condition for MTX withdrawal,[24]. In addition, the randomized clinical design with allocation concealment, the blind evaluation of disease activity status and use of validated RA scores,[28-29] allowed a precise analysis of flares. Moreover, a balanced distributions of demographic profile, disease features and treatment were relevant since these are known factors to influence vaccine immunogenicity and flares,[10-22]. The final small sample size of the study is related to the high rate of refusals to participate and the rigorous exclusion criteria, and is an important limitation of this trial. However, the larger than expected benefit of MTX withdrawal allowed the identification of a significant difference between groups for SC and GMT.

We provide herein novel evidence of an increment of approximately 25% in anti-SARS-CoV-2 antibodies induced by Sinovac-CoronaVac vaccine with temporary MTX withdrawal. Such improvement is very similar to the 20% increase first described regarding MTX discontinuation for 2 weeks after influenza vaccine,[15], and could therefore partially reduce the deleterious effects in seroconversion induced by MTX reported for Sinovac-CoronaVac vaccine,[12] and BNT162b2 mRNA COVID-19,[13, 16]. This immunogenicity enhancement was observed even with a high frequency of combined DMARD therapy and corticosteroids, factors that could further impair immune response to COVID vaccine,[12-13]. Importantly, MTX dose was comparable between the groups and all patients had doses above 10mg/week, in line with the observation that only patients with doses greater than 7.5mg/week benefited from MTX withdrawal after influenza vaccine,[15].

Concerning combination therapy, the distribution of drugs was alike between the groups, equalizing possible additional harmful effects of different DMARD. We also deliberately excluded patients under rituximab, due to well-known effect on humoral immunogenicity and the heterogeneity of phases of treatment cycles,[11-12, 17-21]. In this context, multiple regression analyses revealed that neither combined DMARD nor prednisone impacted the benefit of MTX temporary discontinuation.

Safety related to vaccine and MTX withdrawal intervention was carefully assessed and included several composite measures. Longitudinally, CDAI, SDAI, DAS28-CRP and CRP had similar behaviors between the groups, increasing after the first dose and remaining stable after the second dose. In fact, the increase in disease activity measures, even in MTX-maintain group, is in accordance with the 20% flare rate of SDAI after BNT162b2 mRNA vaccination,[11].

Considering the rate of flares, MTX-hold and MTX-maintain groups were also comparable regarding DAS28-CRP criteria. The similar criteria with DAS28-ESR was used in previous influenza vaccine MTX withdrawal studies,[14-15] and performed better than other DAS28 flare definitions according to OMERACT,[32]. In our trial, however, CDAI>10 showed to be more sensitive than DAS28-CRP, detecting significantly more flares in MTX-hold in comparison to MTX-maintain group. Of note, the magnitude of CDAI variation was similar between the groups.

The Sinovac-CoronaVac vaccine was well tolerated, with no severe side effects. However, MTX-hold group reported a higher frequency of myalgia and vertigo. The former manifestation may be associated with the vaccine or related to underlying disease activity. In conclusion, this is the first randomized study to show a benefit of MTX withdrawal in the immunogenicity of COVID-19 vaccine. In light of these results, we recommend MTX withdrawal for 2 weeks after each vaccine shot in patients with CDAI<10, with close disease activity surveillance. This strategy may be even safer for other vaccines with longer interval between doses or single dose schedules.

## Data Availability

All data produced in the present study are available upon reasonable request to the authors

## Acknowledgements

We thank the contribution of the Central Laboratory Division, Registry Division, Security Division, IT Division, Superintendence, Pharmacy Division and Vaccination Center for their technical support. We also thank the volunteers for participating in the three in-person visits of the protocol and for handling the biological material, and those responsible for the follow-up of all participants.

## Author’s contribution

Araújo, Medeiros-Ribeiro, Saad, Yuki, Aikawa, and Bonfa conceived and designed the study. Araújo, Medeiros-Ribeiro, Saad, Bonfiglioli, Domiciano, Shimabuco, Silva M, Yuki, Pedrosa, Kupa, Zou, Pereira, Silva C, and Aikawa reviewed charts, selected, and invited potential patients for the study. Araújo and Silva M were the 2 non-blinded researchers and were responsible for randomization, enrollment, procedures explaining, informed consent, safety surveillance and patient follow-up by telephone for adherence purposes. Medeiros-Ribeiro, Saad, Bonfiglioli, Domiciano, and Shimabuco were the blinded investigators responsible for disease activity measures.

Kupa, Pedrosa and Bonfa organized and supervised blood collection and vaccination protocol. Pasoto supervised serum processing, SARS-CoV-2-specific antibody ELISA/neutralization assays and SARS-CoV-2 RT–PCR. Medeiros-Ribeiro, Saad, Bonfiglioli, Domiciano, Shimabuco, Yuki, Pasoto, Silva C, Pedrosa, Kupa, Zou, Pereira, Aikawa, and Bonfa participated in data collection and analysis and clinical data management. Araújo, Medeiros-Ribeiro, Saad, Yuki, Pasoto, Silva C, Kupa, Aikawa, and Bonfa were responsible for writing and revision of the manuscript.

All authors helped to edit the manuscript.

