## Supplemental material for "A RANDOMIZED CLINICAL TRIAL OF 2-WEEK METHOTREXATE DISCONTINUATION IN RHEUMATOID ARTHRITIS PATIENTS VACCINATED WITH INACTIVATED SARS-COV-2 VACCINE"

**Supplemental Figure 1 – Date diagram for methotrexate withdrawal guidance**

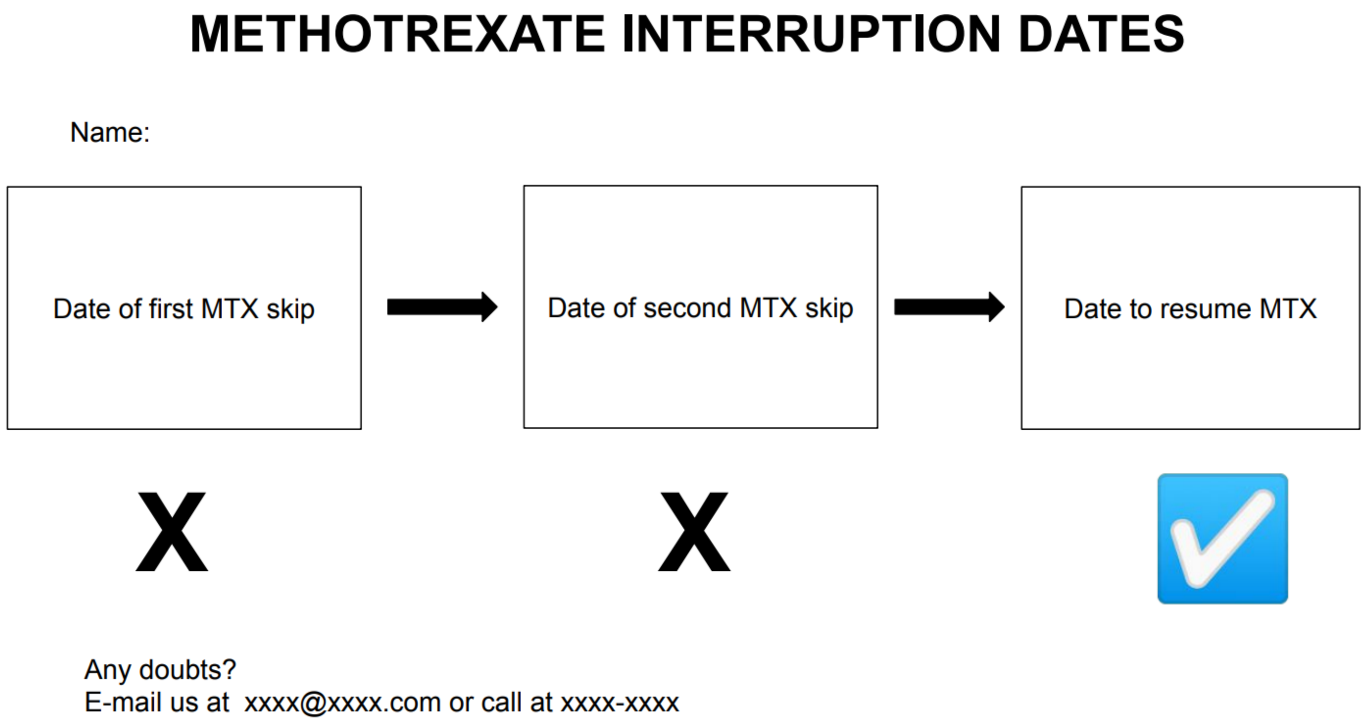

**Supplemental Table 1 – Baseline characteristics of rheumatoid arthritis (RA) patients who finished the study protocol regarding IgG antibodies and NAb after two doses of Sinovac-CoronaVac vaccine (n=92)**

| | Positive IgG after two doses (n=59) | Negative IgG after two doses (n=33) | *p* | Positive NAb after two doses (n=50) | Negative NAb after two doses (n=42) | *p* |
|---|---|---|---|---|---|---|
| Demographics | 55 (42.5-64.5) | 66 (59-69) | 0.001 | 52.5 (40.5-67) | 62.5 (55.3-69) | 0.006 |
| Current age, years | 20 (33.9) | 24 (72.7) | <0.001 | 18 (36) | 26 (61.9) | 0.013 |
| Age >60 years | 53 (89.8) | 31 (93.9) | 0.707 | 47 (94.0) | 37 (88.1) | 0.462 |
| Female sex | 29 (49.2) | 15 (45.5) | 0.733 | 23 (46.0) | 21 (50) | 0.702 |
| Caucasian race | 55 (42.5-64.5) | 66 (59-69) | 0.001 | 52.5 (40.5-67) | 62.5 (55.3-69) | 0.006 |
| Baseline disease activity | | | | | | |
| CDAI | 5.0 (3.0-8.0) | 6.0 (4.0-8.0) | 0.469 | 6.0 (3.0-8.0) | 6.0 (3.0-8.0) | 0.829 |
| SDAI | 6.1 (3.1-9.1) | 7.1 (4.1-9.3) | 0.339 | 6.1 (3.2-9.4) | 7.0 (3.8-9.2) | 0.763 |
| DAS28-CRP | 2.43 (1.82-3.05) | 2.27(2.11-2.90) | 0.843 | 2.45 (1.83-3.10) | 2.33 (1.98-2.97) | 0.742 |
| CRP, mg/dl | 3.3 (1.0-9.2) | 8.9 (7.3-9.8) | 0.154 | 1.6 (0.9-9.2) | 7.6 (3.8-9.8) | 0.161 |
| TJC | 1 (0-2) | 0 (0-1) | 0.086 | 1 (0-2) | 0 (0-1) | 0.259 |
| SJC | 0 (0-1) | 1 (0-1) | 0.297 | 1 (0-1) | 1 (0-1) | 0.727 |
| PGA | 3 (1-4) | 3 (2-4) | 0.491 | 3 (1-4) | 3 (1-4) | 0.570 |
| EGA | 1 (1-2) | 1 (1-2) | 0.223 | 1 (1-2) | 1 (1-2) | 0.766 |
| Current therapy | | | | | | |
| Prednisone | 25 (42.4) | 12 (36.4) | 0.573 | 18 (38) | 19 (45.2) | 0.368 |
| Prednisone dose, mg/day | 5 (2.5-5) | 5 (5-5) | 0.406 | 5 (2.5-5) | 5 (5-5) | 0.278 |
| Withdrew MTX twice | 29 (49.2) | 8 (24.2) | 0.019 | 23 (46) | 14 (33.3) | 0.217 |
| MTX monotherapy | 20 (33.9) | 5 (15.2) | 0.053 | 15 (30) | 10 (23.8) | 0.506 |
| Leflunomide | 8 (13.6) | 11 (33.3) | 0.024 | 7 (14) | 12 (28.6) | 0.086 |
| Sulfassalazine | 1 (1.7) | 0 | >0.999 | 0 | 1 (2.4) | 0.457 |
| Hydroxychloroquine | 11 (18.6) | 6 (18.2) | 0.956 | 12 (24) | 5 (11.9) | 0.137 |
| Tofacitinib | 4 (6.8) | 0 | 0.293 | 3 (6) | 1 (2.4) | 0.623 |
| TNF inhibitor | 11 (18.6) | 7 (21.2) | 0.766 | 12 (24) | 6 (14.3) | 0.242 |
| Abatacept | 6 (10.2) | 7 (21.2) | 0.145 | 4 (8) | 9 (21.4) | 0.066 |
| Tocilizumab | 1 (1.7) | 1 (3.0) | >0.999 | 1 (2) | 1 (2.4) | >0.999 |

Results are expressed in median (interquartile ranges) and n (%). Continuous data were compared using the Mann–Whitney U-test, and categorical variables with the chi-square or Fisher's exact test, as appropriate, as two-sided analyses.

CDAI – Clinical Disease Activity Index; SDAI – Simplified Disease Activity Index; DAS28-CRP - disease activity score with 28 joints and C-reactive protein; CRP – C-reactive protein; TJC – tender joint count; SJC – swollen joint count; PGA – Patient Global Disease Assessment; EGA – Evaluator Global Disease Assessment; MTX – methotrexate; TNF - tumor necrosis factor; Withdrew MTX twice – patients from the MTX-hold group that finished the protocol of withdrawing MTX for 2 weeks after each vaccine dose.

**Supplemental Table 2 - Adverse events after the first and second doses of Sinovac-CoronaVac vaccine in rheumatoid arthritis (RA) patients**

| | After first vaccine dose | | | After second vaccine dose | | |
|---|---|---|---|---|---|---|
| | MTX-hold (n=60) | MTX-maintain (n=69) | *p* | MTX-hold (n=60) | MTX-maintain (n=69) | *p* |
| No symptoms | 29 (48.3) | 29 (42) | 0.473 | 31 (51.7) | 36 (52.2) | 0.954 |
| Local reactions | 13 (21.6) | 13 (18.8) | 0.690 | 7 (11.6) | 7 (10.1) | 0.582 |
| Pain | 9 (15) | 11 (15.9) | 0.883 | 5 (8.3) | 6 (8.7) | 0.941 |
| Erythema | 1 (1.7) | 1 (1.5) | >0.999 | 1 (1.7) | 1 (1.5) | >0.999 |
| Swelling | 3 (5) | 4 (5.8) | >0.999 | 2 (3.3) | 2 (2.9) | >0.999 |
| Bruise | 4 (6.7) | 1 (1.5) | 0.183 | 2 (3.3) | 0 | 0.210 |
| Pruritus | 3 (5) | 1 (1.5) | 0.337 | 0 | 2 (2.9) | 0.499 |
| Induration | 3 (5) | 2 (2.9) | 0.663 | 0 | 2 (2.9) | 0.499 |
| Systemic reactions | 27 (45) | 26 (37.7) | 0.399 | 19 (31.6) | 19 (27.5) | 0.608 |
| Fever | 0 | 2 (2.9) | 0.499 | 2 (3.3) | 3 (4.3) | >0.999 |
| Malaise | 3 (5) | 5 (7.3) | 0.723 | 6 (10) | 4 (5.8) | 0.513 |
| Somnolence | 5 (8.3) | 9 (13) | 0.391 | 4 (6.7) | 3 (4.3) | 0.704 |
| Lack of appetite | 1 (1.7) | 3 (4.4) | 0.625 | 3 (5) | 4 (5.8) | >0.999 |
| Nausea | 2 (3.3) | 4 (5.8) | 0.685 | 4 (6.7) | 3 (4.3) | 0.704 |
| Vomit | 1 (1.7) | 2 (2.9) | >0.999 | 1 (1.7) | 0 | 0.465 |
| Diarrhea | 3 (5) | 3 (4.4) | >0.999 | 4 (6.7) | 1 (1.5) | 0.183 |
| Abdominal pain | 2(3.3) | 4 (5.8) | 0.685 | 4 (6.7) | 1 (1.5) | 0.183 |
| Vertigo | 4 (6.7) | 6 (8.7) | 0.750 | 7 (11.7) | 1 (1.5) | 0.024 |
| Tremor | 3 (5) | 2 (2.9) | >0.999 | 5 (8.3) | 1 (1.5) | 0.096 |
| Headache | 10 (16.7) | 11 (15.9) | 0.657 | 12 (20) | 8 (11.6) | 0.188 |
| Fatigue | 6 (10) | 4 (5.8) | >0.999 | 7 (11.7) | 5 (7.3) | 0.389 |
| Sweating | 3 (5) | 2 (2.9) | 0.663 | 4 (6.7) | 1 (1.5) | 0.183 |
| Myalgia | 5 (8.3) | 4 (5.8) | 0.733 | 10 (16.7) | 3 (4.3) | 0.037 |
| Muscle weakness | 2 (3.3) | 5 (7.3) | 0.448 | 8 (13.3) | 3 (4.3) | 0.112 |
| Arthralgia | 11 (17) | 7 (10.1) | 0.181 | 9 (15) | 5 (7.3) | 0.158 |
| Back pain | 6 (18.3) | 7 (10.1) | 0.978 | 9 (15) | 4 (5.8) | 0.141 |
| Cough | 4 (6.7) | 4 (5.8) | >0.999 | 5 (8.3) | 3 (4.3) | 0.471 |
| Sneezing | 6 (10) | 10 (14.5) | 0.440 | 4 (6.7) | 6 (8.7) | 0.750 |
| Coryza | 3 (5) | 6 (8.7) | 0.502 | 3 (5) | 6 (8.7) | 0.502 |
| Stuffy nose | 3 (5) | 2 (2.9) | >0.999 | 4 (6.7) | 4 (5.8) | >0.999 |
| Sore throat | 2 (3.3) | 6 (8.7) | 0.283 | 3 (5) | 5 (7.3) | 0.723 |
| Shortness of breath | 2 (3.3) | 1 (1.5) | 0.597 | 3 (5) | 0 | 0.094 |
| Conjunctivitis | 0 | 2 (2.9) | 0.499 | 2 (3.3) | 0 | 0.214 |
| Pruritus | 0 | 0 | >0.999 | 1 (1.7) | 0 | 0.465 |
| Skin rash | 1 (2.1) | 2 (3.6) | >0.999 | 0 | 4 (3.6) | 0.123 |

For safety analyses, all patients who adhered to protocol were included. Results are presented in n (%). Categorical variables were analyzed with the chi-square or Fisher's exact test, as appropriate, as two-sided analyses. MTX – methotrexate.
